# Spatially varying effect estimates of mediation in the presence of spatial auto-correlation

**DOI:** 10.64898/2026.10.30.26366937

**Authors:** Kristen Hansen, Yiqun Ma, Armin Schwartzman, Tarik Benmarhnia

## Abstract

The use of mediation analysis is common in epidemiology to decompose an exposure-outcome total effect into direct and indirect effects via a given set of mediators. The counterfactual framework in causal inference allowed for great improvements in mediation analysis to define identification assumptions and clarify sources of bias in different settings. Multiple mediation methods have been proposed to consider confounders of various types and for different settings such as longitudinal designs. However, in some settings we may reasonably expect spatially heterogeneous estimates of both natural direct and indirect effects. In various research areas, especially environmental and social epidemiology, we would expect there to be spatial dependence that could be leveraged. In this paper, we describe and compare a set of approaches to perform spatial mediation analyses and conduct an extensive set of simulations to quantify their performance. We also present a case study assessing the spatially heterogeneous mediating role of PM_2.5_ in the association between income and cardiovascular diseases. We provide all code via GitHub for replication and reproducibility purposes. We find that using a framework combining geographically weighted regression (GWR) and other spatially varying coefficient (SVC) models and the product method of mediation analysis is accurate and precise at estimating spatially heterogeneous effect estimates and handling spatial confounding in a grid setting. Such spatial approaches can be extended to any mediation approach and various confounding structures.

## Introduction

Many epidemiological research questions require more than estimating the total effect of an exposure on a given outcome. Such questions involve, for example, the decomposition of the total effect into separate causal pathways. One common strategy is to break down the total effect into a direct effect of the exposure and an indirect effect of the exposure through one or multiple mediators. Mediation analysis is used frequently and widely in many fields and diverse research contexts. In the field of environmental epidemiology, mediation analysis has recently been used in relation to the role of air pollution on ethnic health disparities (Benmarhnia et al, 2021) and in analyses of psychological outcomes like academic burnout (Ye et al, 2011). Mediation has been used in social epidemiology research on socioeconomic status and mortality (Hossin, et al 2019), an in epidemiological research on blood pressure and tooth loss (Mendes, et al 2021) and cancer prognosis (Syriopoulou, et al 2021).

The most commonly used approach for mediation analysis is based on the product method, proposed by Baron and Kenny (1986). This approach works in special circumstances but is marred by many restrictive assumptions. Over the past decade, mediation analysis methods to assess the importance of different pathways and mechanisms have experienced incredible growth. However, implementing such approaches requires particular identification assumptions (Kaufman 2004). Developments in the methodology of mediation attempt to alleviate required assumptions and broaden the contexts in which mediation can be used. The counterfactual framework has been a large part of this development. A formal counterfactual approach to mediation analysis was developed in 1992 (Robins & Greenland, 1992; Pearl, 2001). It was shown that the total effect can always be decomposed into natural direct and indirect effects, regardless of the underlying statistical model (Lange et al, 2012; Vanderweele et al, 2014). Since then, there has been extensive evolution in the estimation of these effects in time-to-event outcomes (Aalen et al, 2019; Cho et al, 2018) as well as time varying confounders, mediators, and exposures (Lin et al, 2017). In addition, progress has been made to allow for multiple mediators (Bellavia et al, 2017). Furthermore, methodologies have been developed which deal with collider-stratification bias and confounders of the mediator-outcome relationship induced by the exposure (Vanderweele et al, 2014). Finally, recent research has expanded to the use of additional estimators that are robust against misclassification (Wodtke et al, 2020; Rudolph et al, 2018; Rudolph et al, 2020).

The literature handles measured confounding of many types and different modelling techniques. There is however a methodological gap in the literature. Spatial variability in causal inference is beginning to be explored (Reich et al, 2021), but to the best of our knowledge, the literature on mediation analysis does not address spatial heterogeneity, nor does it allow for spatial exposures, mediators, or outcomes. This is especially pertinent for problems relating to environmental or social epidemiology or in any field where data is spatially structured. In addition, mediating relationships can be reasonably expected to vary over space due to confounders and spatial covariance, or due to distribution of the exposures, mediators and population composition. Ignoring spatial relationships when considering mediating relationships may lead to biases due to spatial confounding. In addition, understanding the spatial heterogeneity of direct or indirect effects can help refine targeted interventions in the presence of important spatial heterogeneity of mediated effects.

In this paper, we propose a novel analytical framework to explore spatial heterogeneity in direct and indirect effects. This framework combines the use of regression-based mediation analysis with spatial regression techniques, specifically spatially varying coefficient models. In our simulation study, we explore data contexts of both regular grid data and zip code level data, which is very common in epidemiological research. We investigate different covariance scales, total effect sizes, and varying proportions mediated to account for diverse data contexts. Finally, we include a case study with real data to show how results of this methodological framework could be presented in practice. We provide all code via GitHub (https://github.com/KristenHansen/SpatialMediation) for replication and reproducibility purposes.

## Methods

### Traditional Mediation Analysis

Statistical mediation analysis is used to estimate how an independent exposure variable (*A*) affects an outcome variable (*Y*) through a mediator (*M*). A common approach relies on the product method, which is a regression-based approach. Considering a single mediator, the direct and indirect effects are estimated through the use of three consecutive regression models (Baron et al 1986).

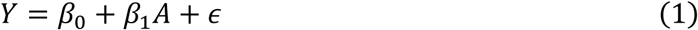

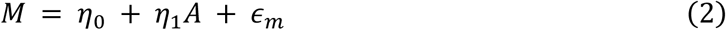

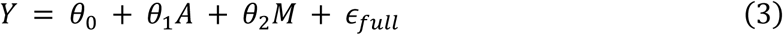

In a linear setting, the product method uses the product of two model coefficients to estimate the indirect effect of *A* on *Y* through *M*. Equation (1) estimates the total effect of *A* on *Y* (*β*_1_). The controlled direct effect is estimated from *θ*_1_ in Equation (3). Finally, the indirect effect is estimated by multiplying *η*_1_ from Equation (2) and *θ*_2_ from Equation (3), with the assumption that *Y* and *M* are both continuous variables. The indirect effect can also be equivalently estimated as the difference of the total effect and the controlled direct effect when all relationships are linear and without interaction.

Recent developments proposed a formal causal framework for mediation analysis using this product method, but these enhancements are applicable to any form of outcome, mediator, and exposure (continuous, binary, counts, etc.) and also allow for exposure-mediator interactions. As with any modeling approach, we assume there is no unmeasured confounding of any of the relationships (exposure-mediator, exposure-outcome, and mediator-outcome). An additional assumption is that there is no confounder (measured or unmeasured) of the mediator-outcome relationship induced by the exposure (*A*). We will henceforth refer to this type of confounding as intermediary confounding. We include more details regarding the notation and estimands of interest in the supplemental materials (see Appendix 1. Counterfactual Framework, Modern Mediation Analysis: Counterfactual Framework). In this paper, we focus primarily on estimating natural direct and indirect effects assuming no exposure-mediator interactions and a single mediator.

### Assumptions and Confounding

We consider four major confounding-based assumptions in mediation analysis (Vanderweele & Vansteelandt, 2010):

1. No unmeasured confounders of the relationship between *A* and *Y*
2. No unmeasured confounders of the relationship between *A* and *M*
3. No unmeasured confounders of the relationship between *M* and *Y*
4. No measured or unmeasured confounders of *M* and *Y* that are affected by *A*

Through use of the counterfactual framework, various weighting and standardization methods have been developed to address assumptions including the fourth and most restrictive assumption. Weighting methods such as Inverse Probability of Treatment Weighting (IPTW), Inverse Odds Ratio Weighting (IORW), and others handle multiple mediators, interactions, and measured intermediary confounding. Additionally, use of G-Computation, a standardization method, can address time-varying confounding, time-to-event outcomes, and exposure induced mediator-outcome confounding and multiple mediators. Stochastic mediation (Rudolph et al., 2018) can allow for exposure induced mediator-outcome confounding and multiple mediators. Thus, there are many methods for handling assumptions in this context that are not available in the spatial domain. While we do not emphasize this source of bias in the present paper, alternative methods that have been proposed can be coupled with the spatial models we introduce below. In this paper, we focus on applying the product method.

### Spatially Varying Coefficient Models

There exist various methods for spatial regression, but they have not been used yet in the context of mediation analyses. Thus, we can extend the use of linear regression in mediation to the spatial domain through the use of spatial regression. We will consider two distinct methods within a single class of models referred to as Spatially Varying Coefficient (SVC) models, which allow model coefficients to vary over space. Specifically, we will focus on Geographically Weighted Regression (GWR) and Moran based SVC (M-SVC) for large data sets. We chose these two methods as they are computationally tractable and analogous to linear modeling with no spatial dependence.

SVC modeling is a popular method for spatial regression in many applied science fields including physics, marine biology, geography, climatology, atmospheric sciences, and urban planning (Paulson & Jiang, 1995; Zang et al, 2021; Wang et al, 2008; Gan et al, 2022; Jiang et al, 2021). As a brief summary, consider that a response variable is sampled at *N* sample sites distributed across a study region. The simple linear SVC has the following form:

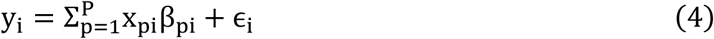

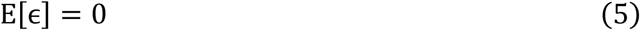

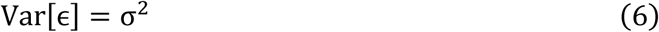

where *x_pi_* represents the *p*-th explanatory variable at the *i*-th sample site and *β_pi_* represents the *k*-th SVC, where *K* is the number of sample sites multiplied by the number of explanatory variables. Finally, *σ*^2^ is a variance parameter. We include more details on these spatial models in the supplemental materials (See Appendix 2. Spatially Varying Coefficients Models: Details on two modeling paradigms).

### Combining Mediation Analysis and SVC modeling

SVC models are analogous to linear regression or mixed effect models, both of which are employed for traditional mediation analysis (Bind et al., 2016). For this reason, under similar assumptions to the product method, we can extend this to a spatial setting. We can fit two SVC models with the forms from Equations (2) and (3) using either GWR or Moran SVC modeling.

From there, the product method can be employed to provide local estimates for Natural Direct Effects (NDE) and Natural Indirect Effects (NIE).

GWR requires the selection of a bandwidth (see details in Appendix 2), for which we will employ a cross-validation (CV) procedure to select for both models (2) and (3), however M-SVC does not require this step. The modeling procedure employs the use of two packages *spmoran* (Murakami) for the M-SVC modeling and *GWModel* for the GWR modeling.

Thus, the procedure has the following form:

1) Choose the bandwidth for the y model of the form of Equation (1). This is required only for GWR models.
2) Fit a model of the form of Equation (1):

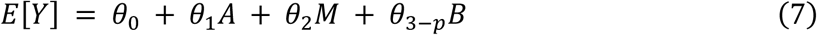

where *A* is the exposure, *M* is the mediator, and *B* represents any confounders.

3) Choose bandwidth for the *M* model (Equation (2)) in GWR.
4) Fit a model of the form of Equation (2),

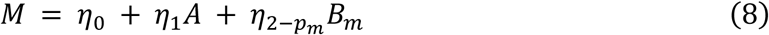

where *B_m_* are the measured confounders of the causal pathway from *A* → *M*.

5) Estimate errors in estimation of *Y* using RMSE (Root Mean Square Error) and squared errors at each location.
6) Estimate spatially varying estimates of NDE and NIE by extracting coefficients from each model and using the procedure outlined in the traditional mediation section to multiply coefficients.
7) Estimate the coverage probability using the standard errors from the SVC models and the true median NDE and NIE values.

There are a few advantages and disadvantages to these two SVC models that are worth mentioning prior to the presentation of our simulation or empirical results. The main difference between the two methods comes in the form of a trade-off. We must choose whether we prefer computational efficiency over estimation accuracy. The M-SVC method is much more computationally efficient as *N* increases due to the approximation of the eigenvector-eigenvalue decomposition. Although under most situations, the data will not be large enough to cause GWR to take longer than an hour. The GWR method has the potential to be more accurate due to no approximation or restrictions on the maximum likelihood estimation. However, GWR does introduce human bias by the choice of spatial covariance function, and it has no adaptive scale parameter, which greatly restricts the contexts in which it can be applied. GWR, although popular due to its implementation in many software platforms, requires particular attention as default settings do not always coincide with the way the data is structured.

### Validation

To validate this method of spatial mediation we will use RMSE to estimate bias, and standard error estimates from the two SVC models to give coverage probability of our NDE and NIE estimates for each setting.

### Simulation Study Design

To test the efficacy of combining the traditional Product method of mediation analysis and spatial regression techniques, M-SVC and GWR, we performed a simulation study in a variety of settings. We wanted to test in different mediation scenarios and across different values of the range parameter for spatial auto-correlation. In Table 1, we present the 12 simulation settings to be considered. We consider scenarios where the total effect is both high and low and when the proportion mediated is high and low, with a distribution across space that has scale (range) that takes on three values. We use the terms range and scale here interchangeably, referring to the extent of the spatial correlation. We consider range as a proportion of the size of the image due to the multiple different spatial domains used in this simulation study: both small and large. We consider range parameters that are small compared to image size (1/20), where the range is about 1/5 of the image size, and when range is high at about 1/2 of the image size. We consider baseline spatial confounders, but no confounders that are unmeasured. We do not consider intermediary confounders for simplicity purposes. We study two data scenarios in addition to our 12 simulation settings. We consider a large regular grid setting, in which we use the shape of California and a grid system that has 0.1 ° × 0.1° cells. Finally, we aggregate the regularly gridded data into ZCTA areas. More details on the simulation settings are presented in Tables S1 and S2.

**Table 1.** Simulation settings considered in the simulation study. All are considered in the small grid, regular grid setting, and aggregated into regions (zip code).

| **Total Effect** | **Proportion Mediated** | **Range Parameter**<br>**(as a proportion of image size)** |
|---|---|---|
| **0.625** | 4% | 1/20 |
| **0.625** | 4% | 1/5 |
| **0.625** | 4% | 1/2 |
| **0.625** | 36% | 1/20 |
| **0.625** | 36% | 1/5 |
| **0.625** | 36% | 1/2 |
| **6.25** | 4% | 1/20 |
| **6.25** | 4% | 1/5 |
| **6.25** | 4% | 1/2 |
| **6.25** | 36% | 1/20 |
| **6.25** | 36% | 1/5 |
| **6.25** | 36% | 1/2 |

## Results

### Regular grid simulation results

In Table 2 are the RMSE values for both modelling frameworks for all the simulation settings as well as ratios for the GWR RMSE over the M-SVC RMSE. In Table S3, we can see that M-SVC takes longer to fit than GWR, which suggests that in a small area setting GWR does not suffer the disadvantage of time.

**Table 2.** Simulation settings with Standardized RMSE from both outcome and mediator models for regular grid simulations.

| **TE** | **Prop Med** | **Scale** | **RMSE GWR** | **RMSE GWR** | **RMSE M-SVC** | **RMSE M-SVC** | **RMSE Ratio** | **RMSE Ratio** |
|  |  | Degrees | Outcome | Mediator | Outcome | Mediator | Outcome | Mediator |
|---|---|---|---|---|---|---|---|---|
| **0.625** | 4% | 0.5 | 0.32 | 0.51 | 0.48 | 0.67 | 0.66 | 0.76 |
| **0.625** | 4% | 2 | 0.36 | 0.25 | 0.25 | 0.32 | 1.44 | 0.78 |
| **0.625** | 4% | 5 | 0.27 | 0.16 | 0.17 | 0.21 | 1.58 | 0.76 |
| **0.625** | 36% | 0.5 | 0.33 | 0.53 | 0.49 | 0.67 | 0.67 | 0.79 |
| **0.625** | 36% | 2 | 0.23 | 0.26 | 0.26 | 0.34 | 0.88 | 0.76 |
| **0.625** | 36% | 5 | 0.30 | 0.15 | 0.16 | 0.21 | 1.87 | 0.71 |
| **6.25** | 4% | 0.5 | 0.5 | 0.36 | 0.51 | 0.49 | 0.67 | 0.76 |
| **6.25** | 4% | 2 | 0.24 | 0.25 | 0.26 | 0.31 | 0.92 | 0.80 |
| **6.25** | 4% | 5 | 0.29 | 0.16 | 0.17 | 0.21 | 1.70 | 0.76 |
| **6.25** | 36% | 0.5 | 0.32 | 0.52 | 0.48 | 0.69 | 0.66 | 0.75 |
| **6.25** | 36% | 2 | 0.41 | 0.25 | 0.26 | 0.34 | 1.57 | 0.73 |
| **6.25** | 36% | 5 | 0.33 | 0.15 | 0.17 | 0.20 | 1.94 | 0.75 |

In this setting, the small total effect scenarios do not result in significantly higher RMSE values. We see the largest RMSE in settings with the smallest scale parameter. The ratio columns show that performance for the M-SVC model is better for the outcome model in settings with a high scale parameter. Overall, the GWR model handles the mediator model efficiently. This suggests that in a setting with very large scale we should consider M-SVC, but in most scenarios GWR performs very well. The NDE and NIE median estimates for each method are in Table 3. Also displayed are the results when running global (i.e., non-spatial) linear models instead of accounting for any spatial autocorrelation. As expected, the global (non-spatial) linear models are inconsistent and most often do not fit the data well. The GWR and M-SVC have similarly good performance in inference. However, in a few settings GWR underperforms compared to M-SVC.

**Table 3.** The estimated NDE and NIE for the models as well as with a global linear model.

| Sim Setting | Global NDE | GWR Median NDE | SVC Median NDE | LM<br>NDE | Global NIE | GWR Median NIE | SVC Median NIE | LM<br>NIE |
|---|---|---|---|---|---|---|---|---|
| 1 | 0.60 | 0.61 | 0.61 | 0.74 | 0.025 | 0.032 | 0.037 | 0.126 |
| 2 | 0.60 | 0.58 | 0.62 | 0.43 | 0.025 | 0.025 | 0.028 | -0.01 |
| 3 | 0.60 | 0.55 | 0.59 | 0.42 | 0.025 | 0.143 | 0.021 | 1.2 |
| 4 | 0.4 | 0.41 | 0.39 | 0.36 | 0.225 | 0.32 | 0.285 | 0.31 |
| 5 | 0.4 | 0.36 | 0.41 | 0.01 | 0.225 | 0.20 | 0.27 | -0.01 |
| 6 | 0.4 | 0.18 | 0.39 | -0.07 | 0.225 | 0.85 | 0.21 | 1.54 |
| 7 | 6 | 5.96 | 6.0 | 5.88 | 0.25 | 0.26 | 0.26 | 0.14 |
| 8 | 6 | 5.96 | 6.01 | 5.69 | 0.25 | 0.31 | 0.27 | 0.49 |
| 9 | 6 | 6.35 | 6.06 | 6.97 | 0.25 | 0.32 | 0.25 | 0.9 |
| 10 | 4 | 4.02 | 3.98 | 4.05 | 2.25 | 2.21 | 2.16 | 2.10 |
| 11 | 4 | 3.81 | 3.97 | 3.28 | 2.25 | 2.15 | 2.09 | 3.15 |
| 12 | 4 | 3.40 | 3.87 | 3.46 | 2.25 | 1.62 | 2.24 | -0.21 |

The coverage probability and standard errors are displayed in Table 4. We can see from this table that the M-SVC models are more consistent and accurate in estimating the NDE and NIE, with some underestimation of NIE in settings 10–12. The coverage probability varies with the GWR model and the standard errors are incredibly high in simulation settings 3 and 8, suggesting that high scale may lead to low accuracy or precision for GWR models, where the bandwidth of the neighborhood is chosen through cross validation. Furthermore, the accuracy of the NDE estimates is shown to be low in the high mediated effect simulations because the coverage probability is so low.

**Table 4.**
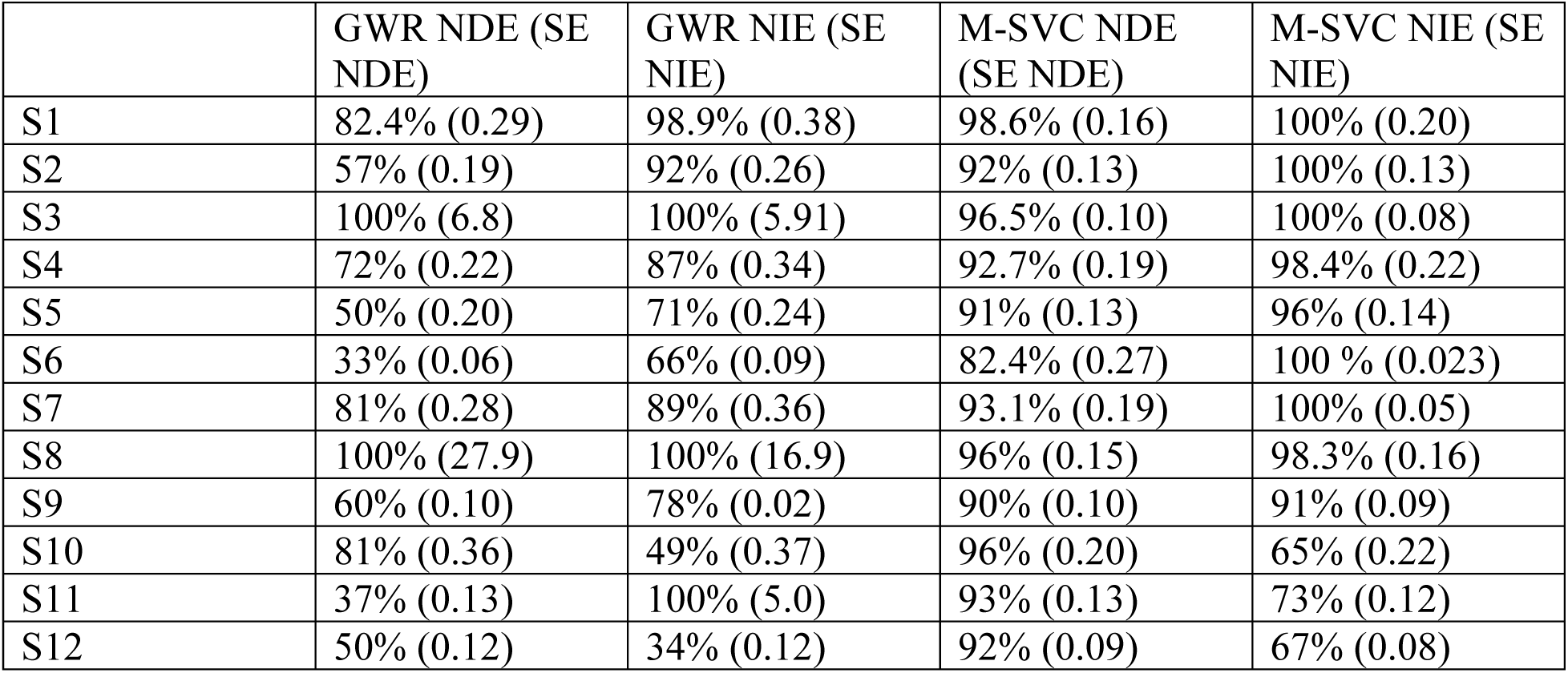
Coverage probability from the GWR and M-SVC models for all 12 simulation settings and both NDE and NIE with standard error estimates from the NDE and NIE estimation.

In Figure 1 and Figure 2 we see the performance of GWR and M-SVC, respectively, in a single instance of simulation scenario 10. The error displayed in Figures 1c and 2c is Gaussian while being low in magnitude. The NDE and NIE for both methods have very similar patterns of differences M-SVC has lower variance in the estimates as well as higher spatial correlation than GWR. Both error plots show roughly a random field, which suggests good model fit in the outcome model. We do not include any confidence intervals or hypothesis tests here as power was not a consideration in this set of simulations. For the low mediated effect simulations, there is not enough power to have statistically significant estimates of NIE.

**Figure 1.**
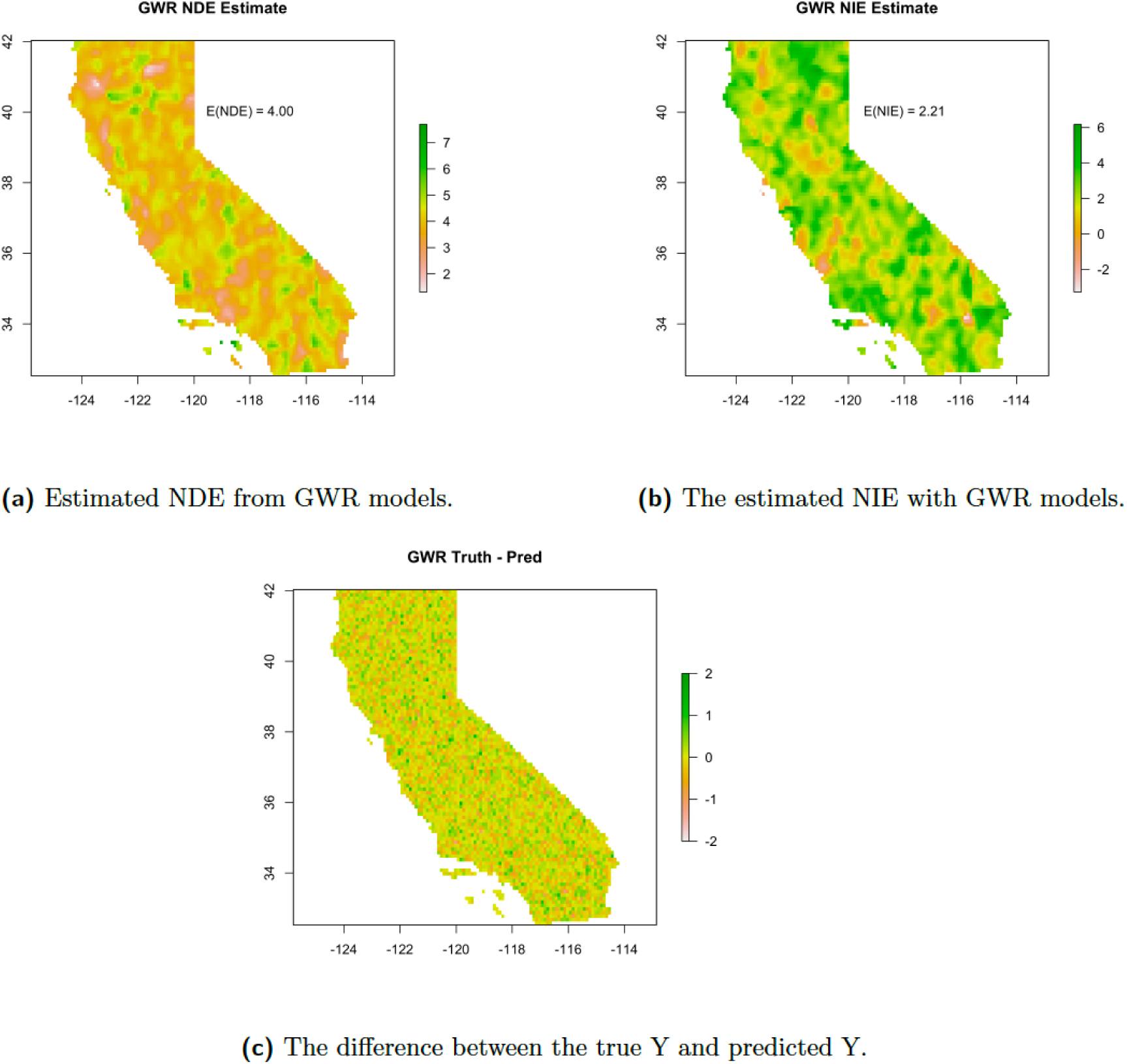
The estimate plots for using GWR in simulation setting 10. We expect the median NDE value to be 4 and the median NIE to be around 2.21.

**Figure 2.**
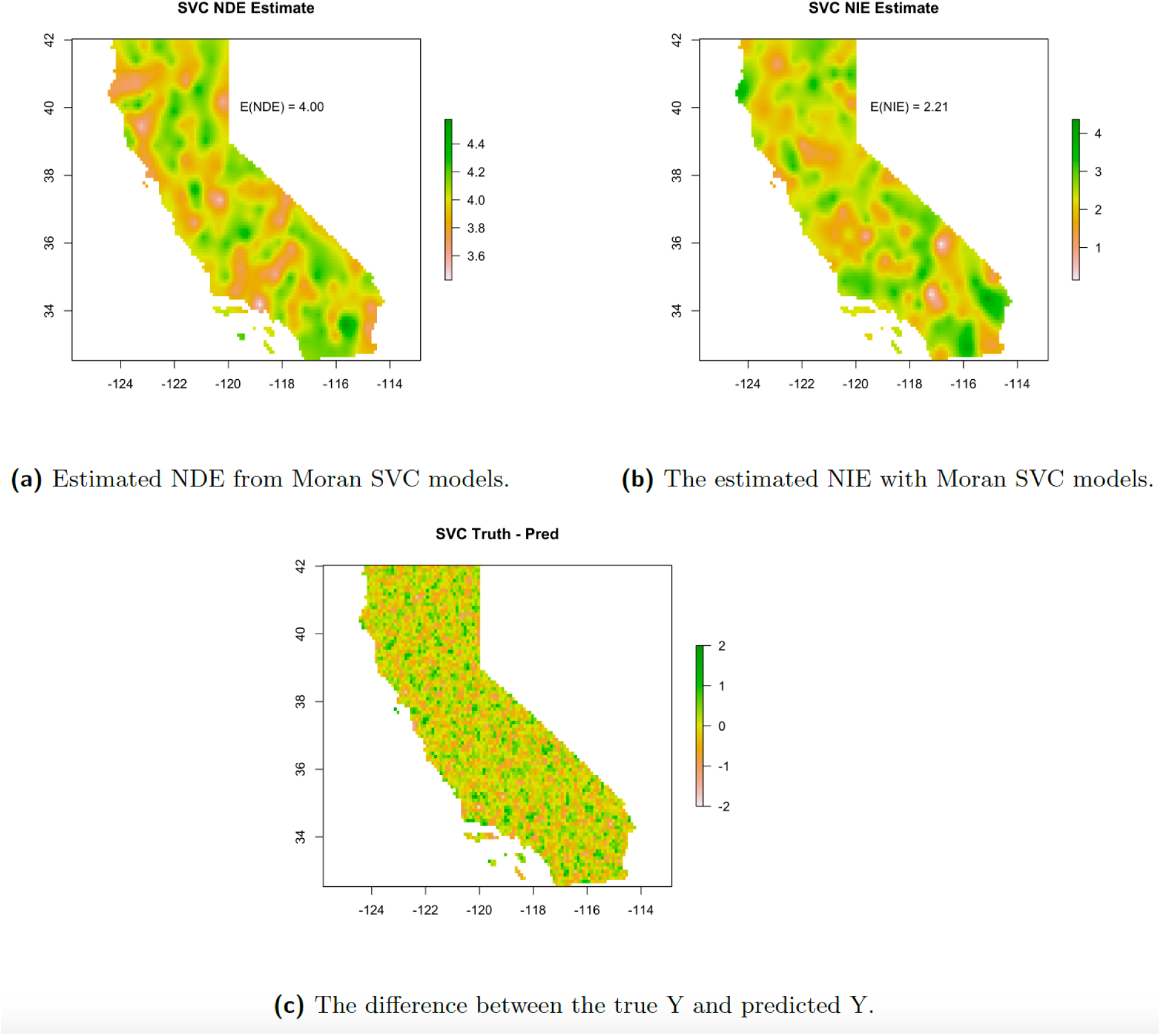
The estimate plots for using Moran SVC in simulation setting 10. We expect the median NDE value to be 4 and the median NIE to be around 2.21.

Whereas M-SVC has a strong advantage, taking only 1.7 minutes on average, GWR takes nearly 6 minutes for a single fit of both the mediator and outcome models without fitting the standard errors (see Table S3). Furthermore, when using a GWR model, one must be careful about the implementation one chooses. In the above grid settings, GWR with fitted standard errors takes about 1.5 hours with the *spgwr* package. We suggest using the *GWmodel* package instead. For this reason and the increased precision of the M-SVC model, we suggest the use of M-SVC.

### Aggregated Areal Region Results

To test the efficacy of this type of analysis on the most common form of spatial data in social or environmental epidemiology, we did the same simulation setting but with data aggregated into ZCTAs in the state of California. This does not represent real data but represents a data situation common where data is aggregated to preserve privacy.

The RMSE values in Table 5 show much larger values than the non-aggregated case in most settings. In particular, when the total effect is low and proportion mediated is low, neither model type converges to the correct values. However, M-SVC has lower RMSE. When the total effect is large, RMSE is similar in value to the non-aggregated case with some bias in the mediator model in simulation setting 7 and 10 suggesting lack of fit when the scale value is too low, which is to be expected because some zip codes are large relative to the scale parameter.

**Table 5.**
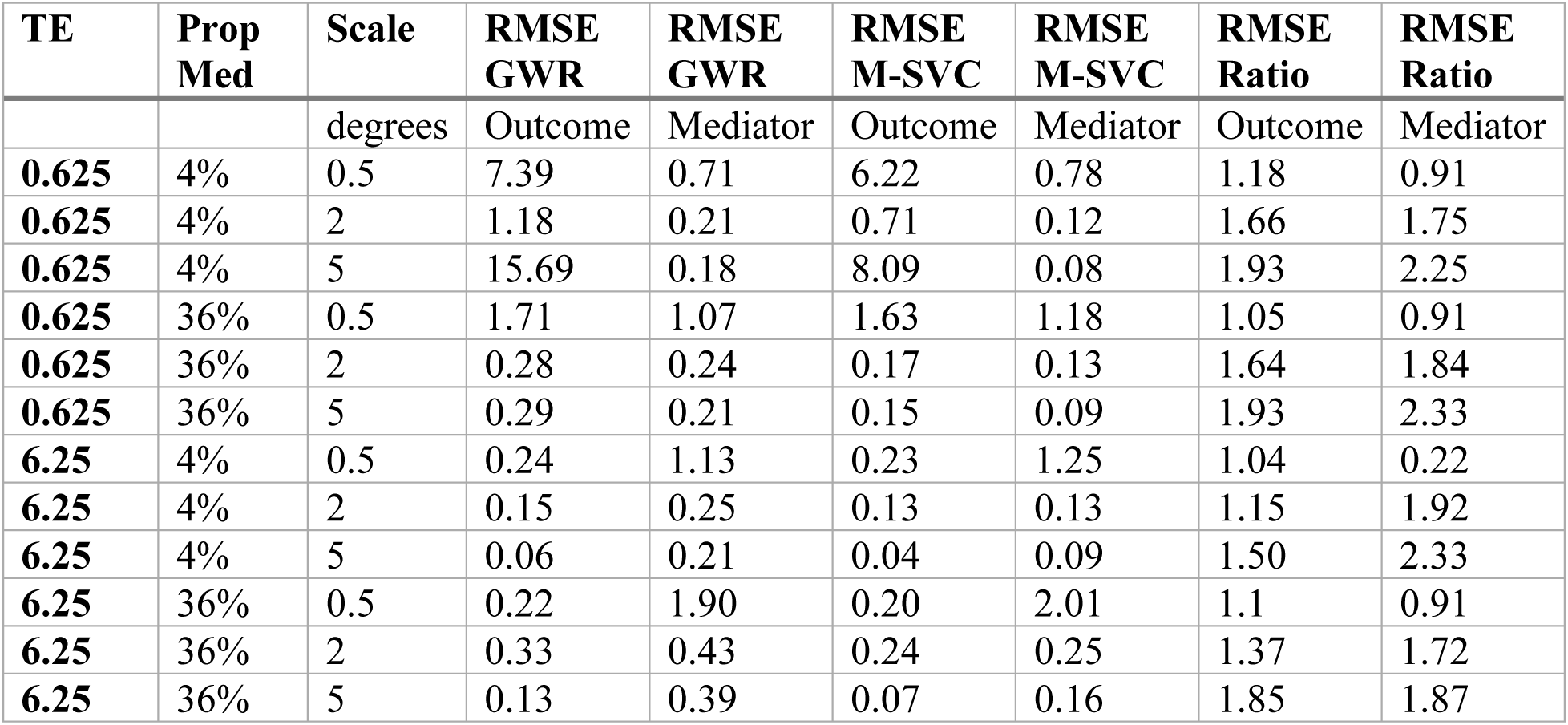
Simulation settings with RMSE from both outcome and mediator models for aggregated simulations for GWR and SVC models.

In Table 6, the NDE and NIE median estimates from both model types are displayed. We can see from the values here that GWR adheres more closely to the true values of NIE but underestimates NDE precipitously. The M-SVC models underestimate both the direct and indirect effects but do a better job in the low total effect settings than GWR. Furthermore, coverage probabilities are not presented here as the standard errors are high in this aggregated setting.

**Table 6.**
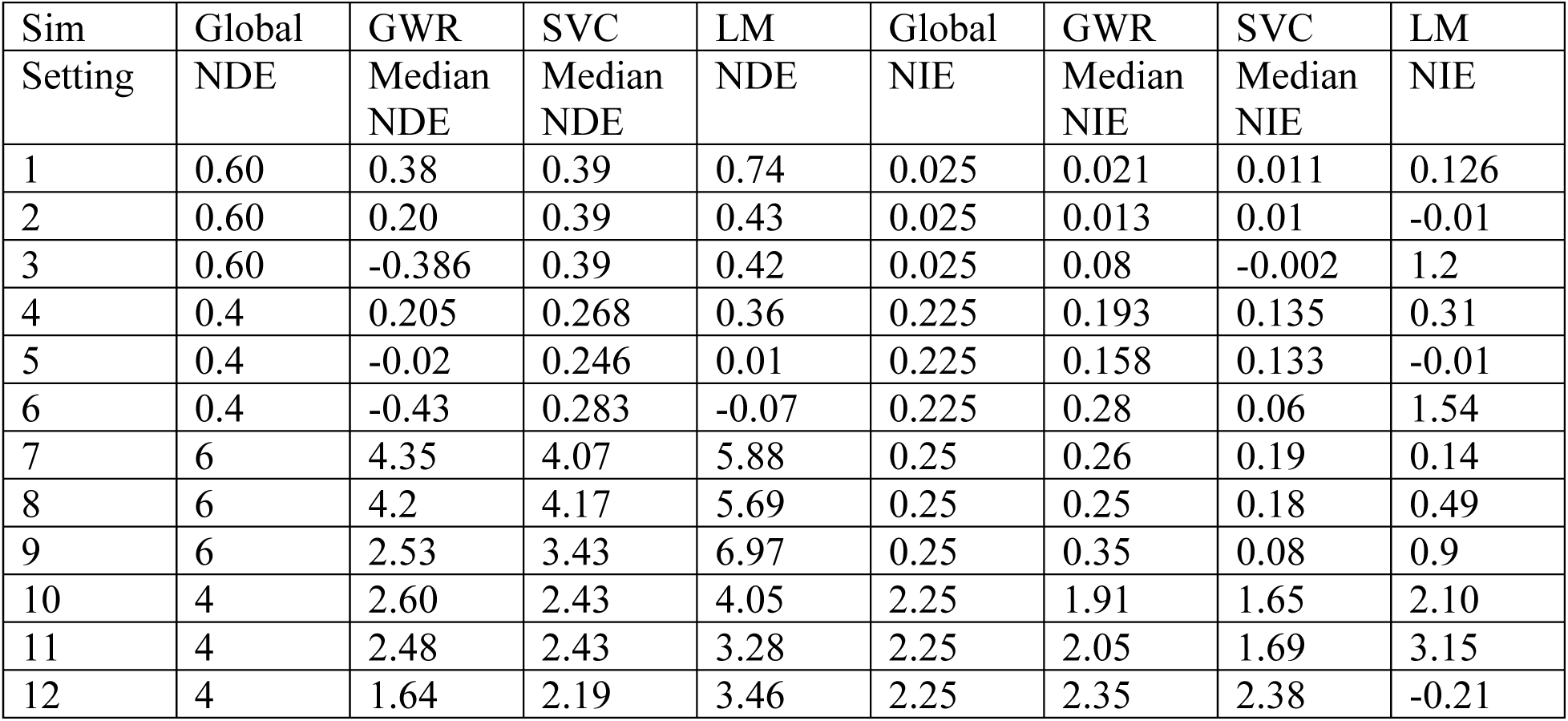
The estimated NDE and NIE for the models as well as with a global linear model.

| Sim | Global | GWR | SVC | LM | Global | GWR | SVC | LM |
|---|---|---|---|---|---|---|---|---|
| Setting | NDE | Median NDE | Median NDE | NDE | NIE | Median NIE | Median NIE | NIE |
| 1 | 0.60 | 0.38 | 0.39 | 0.74 | 0.025 | 0.021 | 0.011 | 0.126 |
| 2 | 0.60 | 0.20 | 0.39 | 0.43 | 0.025 | 0.013 | 0.01 | -0.01 |
| 3 | 0.60 | -0.386 | 0.39 | 0.42 | 0.025 | 0.08 | -0.002 | 1.2 |
| 4 | 0.4 | 0.205 | 0.268 | 0.36 | 0.225 | 0.193 | 0.135 | 0.31 |
| 5 | 0.4 | -0.02 | 0.246 | 0.01 | 0.225 | 0.158 | 0.133 | -0.01 |
| 6 | 0.4 | -0.43 | 0.283 | -0.07 | 0.225 | 0.28 | 0.06 | 1.54 |
| 7 | 6 | 4.35 | 4.07 | 5.88 | 0.25 | 0.26 | 0.19 | 0.14 |
| 8 | 6 | 4.2 | 4.17 | 5.69 | 0.25 | 0.25 | 0.18 | 0.49 |
| 9 | 6 | 2.53 | 3.43 | 6.97 | 0.25 | 0.35 | 0.08 | 0.9 |
| 10 | 4 | 2.60 | 2.43 | 4.05 | 2.25 | 1.91 | 1.65 | 2.10 |
| 11 | 4 | 2.48 | 2.43 | 3.28 | 2.25 | 2.05 | 1.69 | 3.15 |
| 12 | 4 | 1.64 | 2.19 | 3.46 | 2.25 | 2.35 | 2.38 | -0.21 |

Figure 3 and Figure 4 display the NDE and NIE spatial estimates for simulation setting 10. Notably, the NDE is underestimated in both models in this setting and the NIE has some outliers in the GWR fit and is slightly underestimated in the M-SVC fit. The errors in the Y models show higher magnitude error than in the non-aggregated and appear more spatially correlated. However, the errors are generally less than 2 SD from the mean.

**Figure 3.**
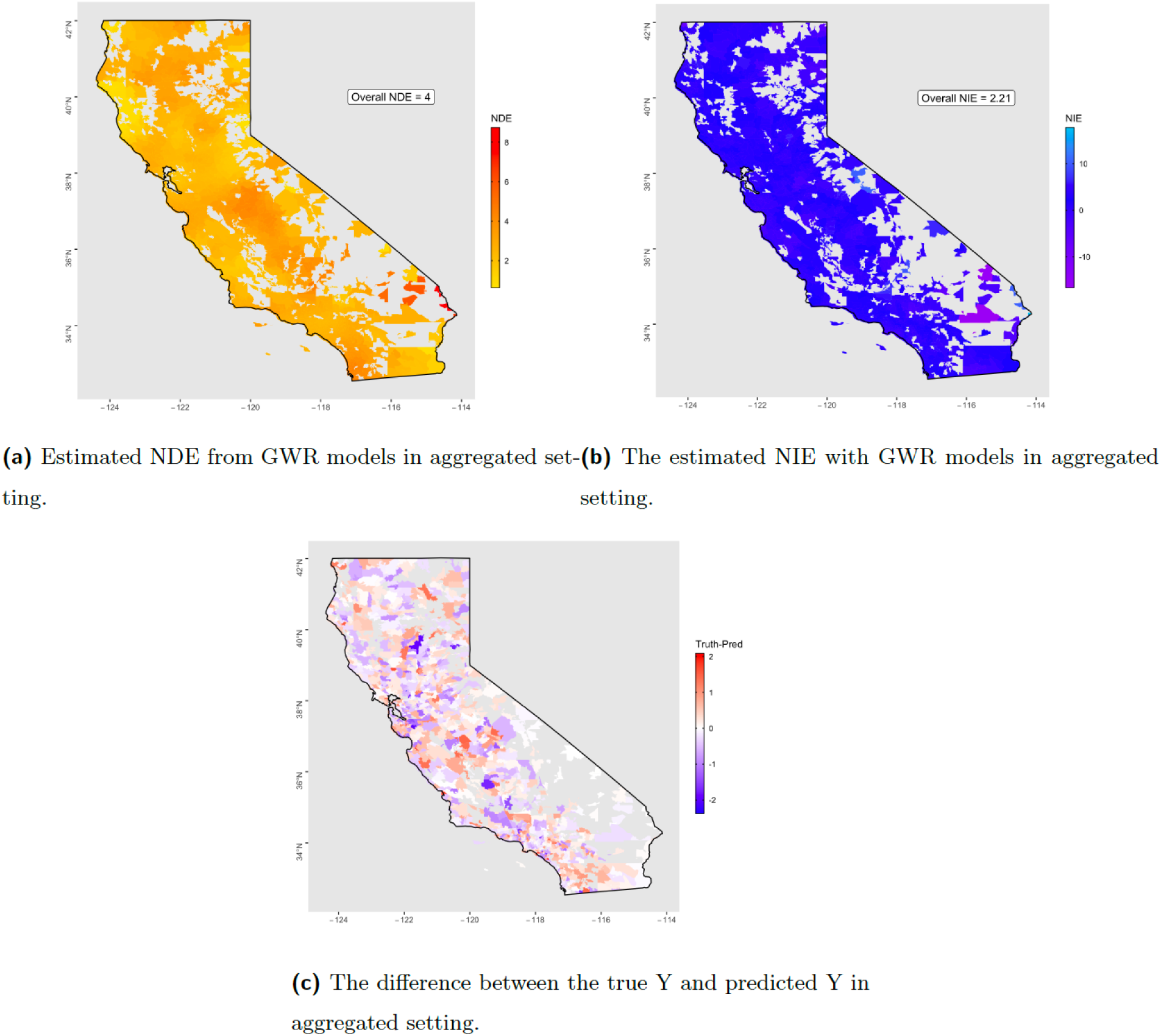
The estimate plots for using GWR in aggregated simulation setting 10. We expect the median NDE value to be 4 and the median NIE to be around 2.21.

**Figure 4.**
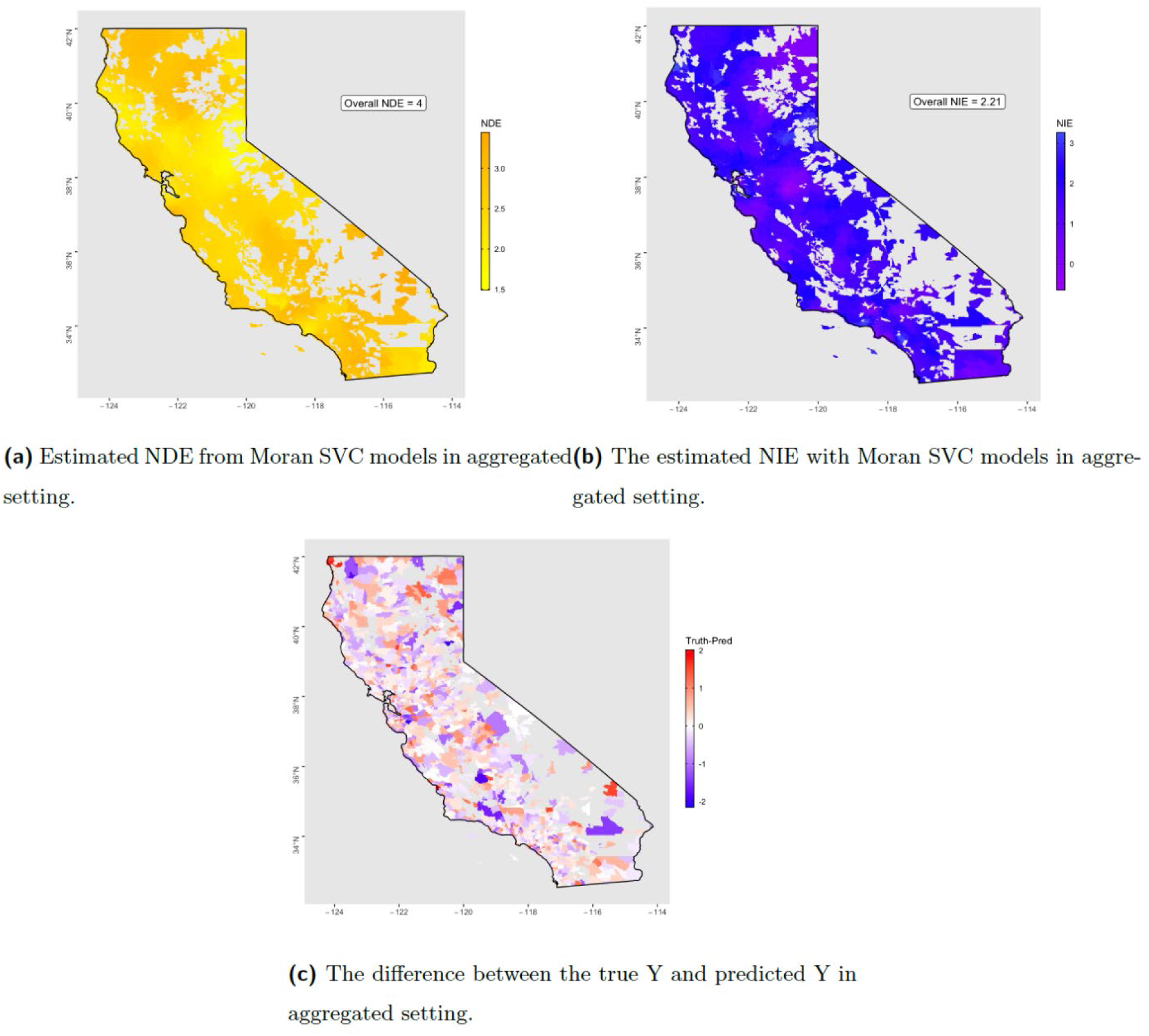
The estimate plots for using Moran SVC in aggregated simulation setting 10. We expect the median NDE value to be 4 and the median NIE to be around 2.21

The results from the aggregated setting show that using zip code level data for geospatial analysis is not optimal. The irregularly spaced regions are not geographic regions, but designations for mail delivery by USPS, leading to vast areas with no coverage and also a low number of neighboring zip codes for many. We see here that GWR and M-SVC models, although they handle the irregularity, have some problems, which we expect to be quite common. Overall though, M-SVC performs better in most settings even though it underestimates the decomposed effect sizes. In general, if zip code level data is the highest resolution available, we recommend including covariates like population, topography information, and area of each zip code to help account for this irregularity.

### Case Study: Modeling Procedure

As a case study, and to illustrate the implementation of a spatial mediation analysis, we use the California Healthy Places Index (HPI) (HPI 2018) and CalEnviroScreen (2019) data to explore the relationship between income and Cardiovascular Disease (CVD) mediated by PM2.5 in Los Angeles County. We evaluate this relationship because it is understood that income can affect health outcomes like rate of CVD (Mosquera 2016), however, pollution like PM 2.5 can modify this relationship as it is also a predictor of CVD (Hystad 2020), but income can affect PM2.5 exposure as well (Finkelstein 2003). The HPI and CalEnviroScreen data is available on the census tract level. This is higher resolution than a zip code and has full coverage of land area, so can lead to better results than the zip code level data. We dichotomize the income variable for simplicity in interpretation by taking the median as the cutoff. We include the education level, proportion white, and employed percentage as confounders in our outcome model. We fit the GWR models for the mediator and the outcome and use the product method to estimate the NDE and NIE.

Figure 5 shows the results from the case study. We can see that the error and bias ranges between -1 and 1. This is a good sign suggesting that the error in the estimation of the outcome falls within one standard deviation of the true value. Bias in general can be due to misattribution of the bandwidth or correlation functions, so if one sees high bias in their outcome models a further exploration of the spatial parameters of the data is suggested.

**Figure 5.**
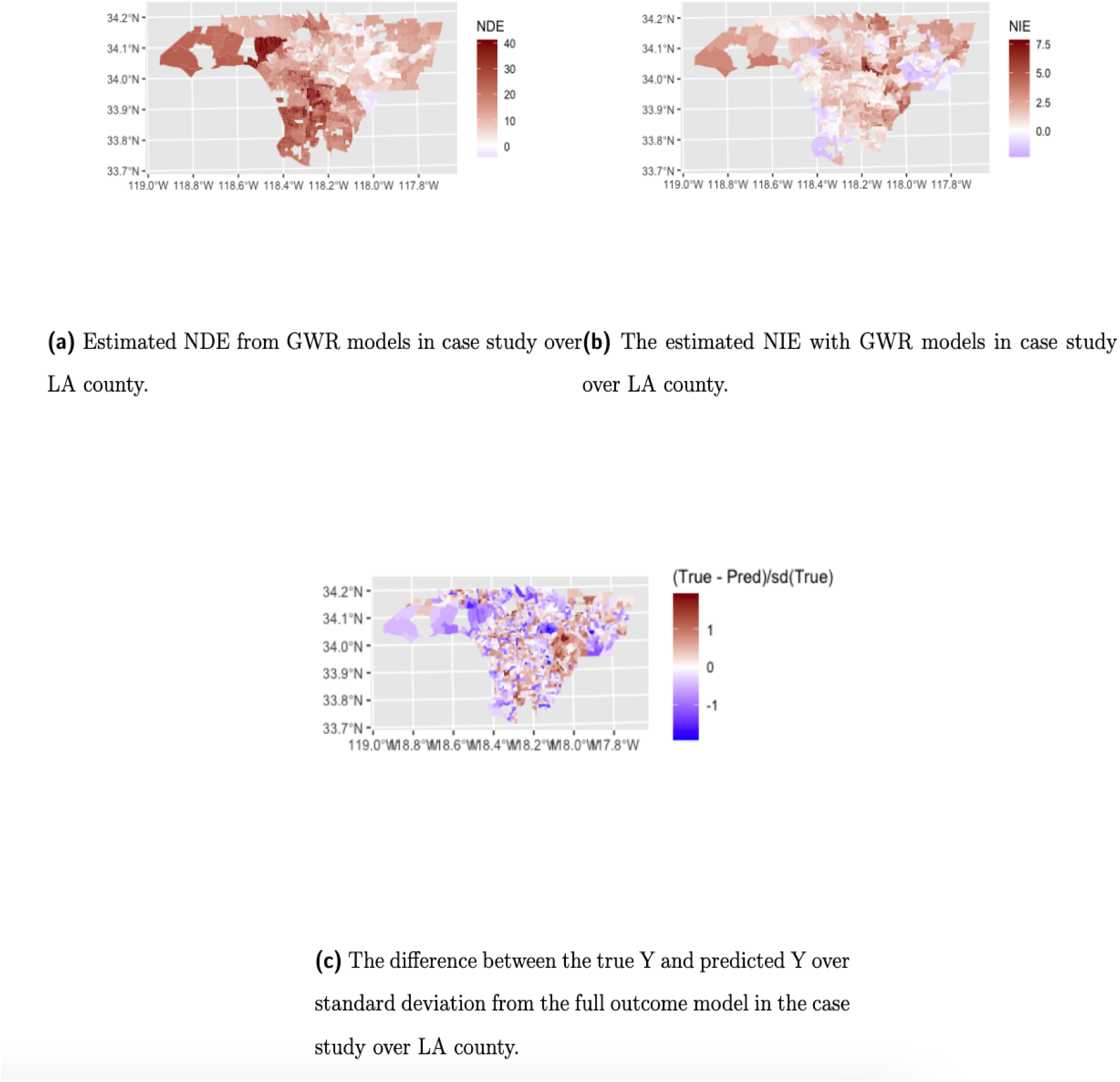
The estimate plots for using GWR with the case study data.

We also see that the mediated proportion ranges from around 5% to 15% here. This suggests that the majority of the total effect of income on CVD percentile is explained by income itself and around 10% mediated by PM_2.5_ exposure. Additionally, in these results we can see that the effect sizes vary over space. The NDE varies from 0 to 40.

## Discussion

In this paper, we propose a novel framework for estimating natural direct and indirect effects of an exposure on a given outcome allowing for spatial heterogeneity. Through simulation, we found that this method of combining traditional mediation analysis and spatial regression is accurate on a gridded dataset.

We find that with the same assumptions as the product method and isotropy we can accurately estimate natural effects on a spatial domain. On a regular grid, the performance of these analysis methods is better than when aggregated at the zip code level. On a gridded dataset in both small and large settings, we see very low bias and good coverage. We find that although both methods have very low bias in most settings, GWR has lower coverage probability and is more inconsistent in performance than M-SVC. M-SVC is thus suggested as the better model framework for this type of data.

For areal unit data, this analysis method is suboptimal at the extremes of total effect, scale, and proportion mediated. In particular, the M-SVC method tends to underestimate effects when the scale variable is low, but in general has lower bias than GWR and lower variability in estimates. GWR can even lead to some negative estimates of NIE and NDE in the small total effect settings. If there is a small total effect and large proportion mediated, M-SVC may be sufficient in a zip-code setting with the inclusion of variables to account for irregularity of each zip code, but this is a narrow window of opportunity for its use. For these reasons, we advise the use of M-SVC based mediation analysis when gridded data is available and GWR with areal units. It is ideal to work in the grid setting in any case as the bias is lower and estimates are more precise.

Both GWR and M-SVC are not designed for areal unit data, and so their performance regarding the areal unit problem should be closely considered. However, these models are continuing to be used with areal units, thus we must find ways to use them that do not violate the assumptions therein. They tend to overestimate the spatial correlation of the data and therefore underestimate the coefficients of the covariates when working with data that are irregularly shaped and spaced. This is a type of spatial confounding. Furthermore, research must be done to adapt a modeling framework that works well for zip code level data without special considerations for large regions rather than a method like this that works well in smaller urban settings but is not ideal for the combination of urban and rural regions, such as an entire state. In principle, we would prefer to work with gridded data. For this reason, research could move into the direction of downscaling epidemiology data to gridded data using various predictors such as census and survey data.

The assumptions under this framework are quite restrictive. In an environmental context we can expect anisotropic data at least for some exposures of interest like air pollution. There are a few possible solutions to this. We can include variables that account for environmental anisotropy in the regression equation, like elevation, topography, and population values as covariates. This, however, will not fully account for anisotropy in all circumstances. We additionally could extend our modeling to the use of anisotropy friendly modeling techniques. For instance, GWR has an extension for adaptive scale parameters (Paez, 2004), and M-SVC could be extended to allow for anisotropic covariance functions. However, anisotropic covariance functions are hard to estimate, so it is not generally done or widely implemented in the field.

In addition to anisotropy this modeling framework cannot estimate accurately in the presence of intermediary confounders. These are the confounders that are affected by the exposure and in turn affect the mediator-outcome pathway. In mediation, this type of confounding can be handled with weighting or standardization techniques like Inverse Probability of Treatment Weighting (IPTW), G Computation, and Stochastic Mediation. Spatial regression with current implementation does not allow for the addition of non-spatial weights along with the geographic weights. Furthermore, G-Computation (even without considering a mediation setting) in a spatial setting has never been done. To allow for G-computation in a spatial setting, there needs to be an extension of time-varying G-computation to two-dimensional space-varying G-computation. This is an area of possible future endeavor.

The limitations discussed are important to consider in the environmental and social epidemiological context as anisotropy and intermediary confounding could be found in the research questions in this field. For that reason, future work will need to address these concerns. However, this method for estimating spatial heterogeneity in direct and indirect effects of an exposure is an essential step in spatial causal inference for environmental and social epidemiology. Furthermore, this method allows investigators to explore research questions where spatially heterogeneous effect decomposition can be expected.

## Data Availability

All data produced in the present work are contained in the manuscript.

https://github.com/KristenHansen/SpatialMediation

## Acknowledgments

We would like to thank Maren Hale for her help in revising this manuscript.

