## Appendix for "Spatially varying effect estimates of mediation in the presence of spatial auto-correlation"

(1) Herbert Wertheim School of Public Health, University of California San Diego, La Jolla, CA, USA
(2) Axle Research and Technology, Rockville, MD, USA
(3) Scripps Institution of Oceanography, University of California San Diego, La Jolla, CA, USA
(4) Irset Institut de Recherche en Santé, Environnement et Travail, UMR-S 1085, Inserm, University of Rennes, EHESP, Rennes, France.

### *Appendix 1. Modern mediation analysis under the counterfactual framework*

Modern mediation analysis has built upon the product method using causal inference literature and counterfactual frameworks (Rubin et al 2005). The counterfactual framework provides notation for fixing relevant variables in non-randomized studies (Rubin 1974). For further details on this notation see *Causal inference: For statistics, social and biomedical sciences: an introduction* by Imbens and Rubin (2015).

The individual causal effect of an exposure on an outcome is the hypothetical contrast between the outcome under the exposure and in the absence of the exposure. The notation for this includes $Y_a$, which is the potential outcome under the exposure, $A = a$. Then $Y_{a^*}$ is the potential outcome under the counterfactual exposure, $A = a^*$ where $a = 1 - a^*$ when there is a binary exposure. It is only possible to observe either $Y_a$ or $Y_{a^*}$. This is known as the fundamental problem of causal inference (Holland 1987). Then the causal effect is defined as $E(Y_a - Y_{a^*})$ (Richiardi et al 2013; Rubin 1974).

In mediation analysis specifically, this framework defines natural direct and indirect effects that sum to the total effect (Pearl 2001). Here $Y_{a,m}$ is the potential outcome under exposure $A = a$ and mediator $M = m$. The natural direct effect is the difference between the outcome if exposed to $A = a$ and the value of the counterfactual outcome if the same realization of all confounders, were exposed instead to $A = a^*$ and the mediator set to the value it would take with exposure $A = a^*$. The natural direct effect is thus defined as $E\left(Y_{a,M(a^*)} - Y_{a^*,M(a^*)}\right)$. It is also possible to estimate controlled direct effects (which is equivalent to natural direct effects in the absence of exposure-mediator interactions), but in this paper we focus only on natural direct effects as our estimand of interest. Finally, the natural indirect effect is defined as $E\left(Y_{a,M(a)} - Y_{a,M(a^*)}\right)$. This is the difference in the outcome if exposed to $A = a$ and $M = M(a)$ and the value of the counterfactual outcome, if exposed to $A = a$ but $M = M(a^*)$ takes on the value $M$ would take if the exposure was $A = a^*$, under the same realization of all confounders under $A = a$.

### *Appendix 2. Spatially Varying Coefficients Models: Details on two modeling paradigms*

#### *Geographically Weighted Regression*

GWR extends Ordinary Least Squares (OLS) linear regression models by accounting for spatial structure and estimates a separate model and local parameter estimates for each geographic location contained in a data set. This local model is based on a subset of the data around the location or the neighborhood of the point. These points are utilized by adopting a differential weighting scheme. Introduced by Brunsdon et al (1998), this method of modeling is very similar to kernel regression (Ruppert et al 1995) in that we have a kernel function. However, in GWR the kernel is not based on similarities in predictor variables, but is instead spatial, considering physical distance.

GWR can be considered a kernel-weighted method of regression. In any kernel regression, there are two major choices to be made. We must choose a kernel function and choose a bandwidth for our "circle of inclusion".

#### *Kernel Function*

The regression model centered at each $x_i$ can be thought of as a weighted OLS regression where the weights for points within the neighborhood follow a particular kernel function and those outside the neighborhood are weighted at 0. The simplest case of a kernel function gives uniform weights to all observations within the neighborhood:

$$w_{ik} = \begin{cases} 1 & d_{ik} < h \\ 0 & otherwise \end{cases} \tag{1}$$

Where $h$ is the radius of the neighborhood and $d_{ik}$ is the distance from a point $x_k$ to the point $x_i$. This weighting is overly simplistic and restrictive as a step function. Continuous kernel functions are more common. A kernel function, *K(d)*, which is a function of distance, should have the following properties (Brunson et al 1998):

1) $K(0) = 1$
2) $\lim_{d \to \infty} K(d) = 0$
3) K is a monotone decreasing function for positive real numbers

The kernel function we will consider for our analysis and simulation will be the Gaussian decay kernel:

$$w_{ik} = e^{-d_{ik}^2/(2h^2)} \tag{2}$$

The Gaussian decay kernel allows for the weight to gradually decay with distance. The constant bandwidth, $h$, in a continuous decay function only provides a measure of the rapidity of decay. Using this weighting scheme, each resulting β estimate in model ( 4 ) will be continuous on $R^2$.

There are several other common Kernel functions including the Exponential function:

$$w_{ik} = e^{-d_{ik}/(h)}$$

and the bi-square function:

$$w_{ik} = \begin{cases} 1 - \left((d_{ik})^2/(2h^2)\right)^2 & d_{ik} < h \\ 0 & Otherwise \end{cases}$$

Ideally, the choice of a Kernel function will depend on the spatial covariance of the observed data. However, in our experience, the bandwidth is more important than the shape of the kernel. There are many approaches to choosing a kernel function apart from the spatial covariance but a Gaussian kernel is the most commonly used in the literature for continuous data (Lu et al 2011).

### *Choosing the Bandwidth*

Choosing the bandwidth is a more important and influential choice in our experience. There are two common ways to choose the bandwidth $h$. The first uses a cross validation method to choose the optimal fixed bandwidth. Here we find the $h$ that minimizes the sum of squared errors at all locations, $i$:

$$\text{CV} = \Sigma_i [y_i - \widehat{y_{\neq \imath}}(\beta)]^2$$

Where $\widehat{y_{\neq \imath}}(\beta)$ is the fitted value of $y_i$ with the observation for the $i^{th}$ site omitted from the calibration. This method is the method we will use when choosing a bandwidth. However, it should be noted that leave one out cross validation can change the spatial dependence structure in an areal data setting. There is an additional method for bandwidth choice that minimizes the Akaike Information Criterion (AIC), but this involves the computation of $n \times n$ matrices where $n$ is the number of sample sites and therefore is not ideal for large spatial datasets.

### *Computation*

Having chosen a weighting function and bandwidth, our β estimates can be computed with the following:

$$\beta = (X^T W X)^{-1} X^T W y \qquad (3)$$

The matrix $W_i$ is the diagonal matrix of the $w_{ik}$ weights:

$$W_i = \begin{pmatrix} w_{i1} & \cdots & 0 \\ \vdots & \ddots & \vdots \\ 0 & \cdots & w_{in} \end{pmatrix}$$

( 4 )

The β in Equation ( 7 ) is a matrix containing all $\beta_{ij}$ for each sample site $i$ and predictor $j$ (from 1 to p). This is the main output from GWR. A row corresponds to all the β estimates for each location. The matrix of β's can be extracted as an $n \times p$ matrix where $p$ is the number of predictors.

### *Moran Spatially Varying Coefficient Model*

Moran SVC (M-SVC) is an approach developed and based on the Moran coefficient (Moran 1950). This coefficient is a diagnostic statistic of spatial dependence. MC when $\boldsymbol{y} = [y(s_1), \dots, y(s_N)]'$ is:

$$\mathrm{MC}(y) = \frac{N}{\mathbf{1'D1}} \frac{\boldsymbol{y}'\boldsymbol{C}\boldsymbol{D}\boldsymbol{C}\boldsymbol{y}}{\boldsymbol{y}'C\boldsymbol{y}}$$

( 5 )

$D$ is a symmetric spatial proximity matrix with 0 diagonals and $C$ is a centering matrix of the form $C = \boldsymbol{I} - \mathbf{11}'/N$, and $N$ is the number of sample sites. When the $\boldsymbol{y}$s are positively spatially dependent values then $\mathrm{MC(y)} > \frac{-1}{N-\mathbf{1}}$. And when negatively dependent, $\mathrm{MC} < \frac{-1}{N-1}$ (Griffith 2003). $D$ is a proximity matrix where the $(i,j)^{\text{th}}$ element is defined by $e^{-d(s_{ij})/r}$ where the range parameter $r$ is given by the maximum distance in the minimum spanning tree connecting all sample sites (Dray et al 2006; Murakami et al 2015).

### *Modeling*

Moran eigenvector SVC (M-SVC) is a modeling approach (Griffith 2008), which is an extension of the Moran eigenvector spatial filtering approach (Griffith 2014). While instability of this approach was alluded to by Helbich and Griffith (2016) and Oshan and Fotheringham (2018), Murakami et al. (2017) extended it to approximate the Bayesian-SVC model, and Murakami et al. (2017; 2018) demonstrated its estimation accuracy through Monte Carlo experiments with a fast computation approach introduced in 2019 (Murakami 2019).

Let $V = CDC$, take $\boldsymbol{E} = [e_1, \dots, e_L]$ to be an $N \times L$ (< $N$) matrix of eigen vectors of the CDC matrix. Furthermore, $\boldsymbol{\Lambda}$ is the diagonal $L \times L$ matrix of the $L$ leading eigenvalues of V $\{\lambda_1, \dots, \lambda_L\}$, these explain the highest proportion of spatial dependence. This method applies $L$ eigen-pairs corresponding to positive eigenvalues from $\boldsymbol{E}$ and $\Lambda$. This method like other SVCs assumes positive spatial dependence by only taking $L$ eigen-pairs corresponding to positive eigenvalues.

The M-SVC coefficients are formulated with a fixed and varying portion with an eigenvector Moran Coefficient (MC) based model of spatial dependence. The overall model for an outcome, $Y$, is as follows:

$$\boldsymbol{y} = b_1\mathbf{1} + \Sigma_{k=2}^{K}\boldsymbol{x}_k \circ \boldsymbol{\beta}_k + \boldsymbol{E\gamma_1} + \boldsymbol{\varepsilon}, \boldsymbol{\gamma_1} \sim N(0, \tau_1^2\Gamma^{\alpha_1}),\ \varepsilon \sim N(0, \sigma^2\boldsymbol{I}) \tag{6}$$

Where $\mathbf{y}$ $(N \times 1)$ is a vector of response variable values at each sample site and $x_k$ $(N \times 1)$ is the vector of the $k^{th}$ covariate for all the spatial sample points, *i.* $\boldsymbol{E\gamma_1}$ captures residual spatial dependence, Murakami and Griffith (2018) showed that the term $\boldsymbol{E\gamma_1}$ with $L$ = 200 greatly reduces residual spatial dependence.

The $\beta_k$ is made from two components, $\beta_k = b_k + \boldsymbol{E\gamma_k}$, where $b_k$ is a constant mean and $\boldsymbol{E\gamma_k}$ is the spatially varying component, composed of eigenvectors and random coefficients, $\boldsymbol{\gamma_k} \sim N(0, \tau_k^2\Gamma^{\alpha_k})$. $\boldsymbol{E\gamma_k}$ has 2 properties: it is interpretable in terms of the Moran Coefficient and the eigenvectors have zero means. Both properties are important to make the $\boldsymbol{E\gamma_k}$ values identifiable and model the spatial dependence.

The $\boldsymbol{\gamma_k}$ vector is a vector of random coefficients depending on two shrinkage parameters, $\tau_k^2$ and $\alpha_k$. The $\tau$ parameter controls the variance of the spatial variation of $\boldsymbol{\beta_k}$**.** The $\alpha$ parameter guides the spatial scale, thus estimating the spatial scale parameter of the SVC rather than using a range parameter, which is treated as fixed in a GWR model. This accelerates model estimation and estimation error remains small (Murukami and Griffith 2018).

We additionally have an error term, $\epsilon$, which we assume is Gaussian with mean 0 and variance $\sigma^2$, that captures non-spatial variance. We assume all spatial variation is captured in the other terms of the model.

### *Parameter Estimation*

The M-SVC model is estimated by the Type II restricted likelihood (empirical Bayes) method that maximizes the log-likelihood of the shrinkage parameters $\Theta = \theta_1 \ldots \theta_k$, where $\theta_k = \{\tau_k^2, \alpha_k\}$ and the log-likelihood of $y$ conditional on $b$ and $\Theta$. If the probability density functions for $y$ and the random coefficients $u$ are Gaussian, the log-likelihoods have an analytical expression. Using this restriction, the parameters can be estimated (Bates 2010). Thus, since $\boldsymbol{u}$ represents the random coefficients, $\mathbf{V}(\boldsymbol{\theta_k})\boldsymbol{u_k} = \boldsymbol{\gamma_k}$ as defined previously.

$$loglik(\Theta) = -\frac{1}{2}\ln|\boldsymbol{P}| - \frac{N-K}{2}\left(1 + ln\left(\frac{2\pi d(\Theta)}{N-K}\right)\right) \tag{7}$$

$$P = \begin{bmatrix} X'X & X'E_1V(\theta_1) & \dots & X'E_kV(\theta_k) \\ X'E'_1V(\theta_1) & V(\theta_1)E'_1E_1V(\theta_1) + I & \dots & V(\theta_1)E'_1E_kV(\theta_k) \\ \vdots & \vdots & \ddots & \vdots \\ V(\theta_k)E'_kX & V(\theta_k)E'_kE_1V(\theta_1) & \dots & V(\theta_k)E'_kE_kV(\theta_k) + I \end{bmatrix}$$

The derivative $\mathbf{d}(\boldsymbol{\theta})$ is equal to:

$$d(\theta) = \left\| y - X\hat{\boldsymbol{b}} - \sum_{k=1}^{K} \boldsymbol{E_k V}(\boldsymbol{\theta_k})\widehat{\boldsymbol{u_k}} \right\|^2 + \sum_{k=1}^{K} \|\widehat{\boldsymbol{u_k}}\|^2$$

The $b$ and the $u$ values are estimated by Equation ( 12 ).

$$\begin{bmatrix} \hat{\boldsymbol{b}} \\ \widehat{\boldsymbol{u_1}} \\ \vdots \\ \widehat{\boldsymbol{u_k}} \end{bmatrix} = \boldsymbol{p}^{-1} \begin{bmatrix} \boldsymbol{X'y} \\ \mathbf{V(\theta_1)E'_1y} \\ \vdots \\ \boldsymbol{V}(\boldsymbol{\theta_k})\boldsymbol{E'_k y} \end{bmatrix} \quad (8)$$

This combination balances the accuracy and complexity of the model. The values for Θ can be estimated by maximizing ( 11 ). This is followed by the estimation of the $\{b, u_1, \ldots, u_K\}$ with Equation ( 12 ). The SVCs are estimated by Equation ( 13 ).

$$\boldsymbol{\beta_k} = \boldsymbol{b_k} + \boldsymbol{E\gamma_k} = \widehat{\boldsymbol{b_k}}\mathbf{1} + \boldsymbol{EV}(\widehat{\boldsymbol{\theta_k}})\widehat{\boldsymbol{u_k}} \quad (9)$$

Finally the residual variance must be estimated as $\widehat{\boldsymbol{\sigma^2}} = \widehat{\frac{\mathbf{1}}{\boldsymbol{N-K}}}\hat{\boldsymbol{\varepsilon}}'\hat{\boldsymbol{\varepsilon}}$ where $\hat{\varepsilon} = y - \widehat{\boldsymbol{b_1}}\,\mathbf{1} - \sum_{k=2}^{K} \boldsymbol{x_k} \circ \boldsymbol{\beta}_k - \boldsymbol{E\gamma_1}$ as defined in equation ( 10 ). The variance of the $\hat{\beta}$estimates are used to evaluate statistical significance. This variance is estimated analogously to a linear mixed effects model.

Likelihood maximization is computationally inefficient especially due to the high number of parameters that must be estimated under a high number of predictors, $K$. Thus, matrix tricks can be employed to speed the processing time of this type of model (Murakami 2019). For the purposes of computational efficiency, there is a fast M-SVC (Murakami 2019). Using the Nystrom extension (Drineas 2005) based approximation of the Moran eigenvectors and eigenvalues (Murakami 2018), this avoids the high computational cost of eigenvalue decomposition. And instead of using every sample point for the development of the proximity matrix a C matrix is computed using $L$ knots where $L \ll N$. $L = 200$ is sufficient to model spatial dependence (Murakami 2018).

### *Small grid simulation results*

In Table 6,we can see the RMSE for the outcome and mediator models in the small 20x20 grid simulation. When our total effect is low, universally the RMSE in the mediator model is much higher than the RMSE for the outcome model. When total effect is high and proportion mediated is high we have the most favorable results of the models with lowest bias. However, in the low total effect and high proportion mediated case, when the scale parameter is high the mediator models fail in both GWR and M-SVC. However, a scale parameter this large is not expected in practice and with a spatial covariance with such a large relative scale, a global linear model may perform just as well. In addition, both modeling techniques perform about equally here. We do not present the standard errors and coverage probability because this setting is under powered, and thus the standard errors are larger than the effect sizes.

**Table S1 Small simulation RMSE results for both GWR and M-SVC**

| **TE** | **Prop Med** | **Scale** | **RMSE GWR** | **RMSE GWR** | **RMSE M-SVC** | **RMSE M-SVC** | **RMSE Ratio** | **RMSE Ratio** |
|---|---|---|---|---|---|---|---|---|
| | | Pixels | Outcome | Mediator | Outcome | Mediator | Outcome | Mediator |
| **0.625** | 4% | 1 | 0.74 | 1.27 | 0.66 | 1.23 | 1.12 | 1.03 |
| **0.625** | 4% | 3 | 0.24 | 0.68 | 0.26 | 0.86 | 0.92 | 0.79 |
| **0.625** | 4% | 10 | 0.10 | 0.34 | 0.57 | 0.34 | 0.14 | 1.0 |
| **0.625** | 36% | 1 | 0.48 | 1.0 | 0.46 | 0.99 | 1.04 | 1.01 |
| **0.625** | 36% | 3 | 0.34 | 0.73 | 0.41 | 0.78 | 0.83 | 0.93 |
| **0.625** | 36% | 10 | 0.56 | 16.6 | 0.43 | 11.3 | 1.3 | 1.46 |
| **6.25** | 4% | 1 | 0.14 | 0.84 | 0.48 | 0.84 | 0.29 | 1.0 |
| **6.25** | 4% | 3 | 0.11 | 0.51 | 0.41 | 0.54 | 0.26 | 0.94 |
| **6.25** | 4% | 10 | 0.02 | 0.27 | 0.03 | 0.20 | 0.66 | 1.35 |
| **6.25** | 36% | 1 | 0.19 | 0.82 | 0.38 | 0.82 | 0.5 | 1.0 |
| **6.25** | 36% | 3 | 0.05 | 0.31 | 0.11 | 0.36 | 0.45 | 0.86 |
| **6.25** | 36% | 10 | 0.27 | 0.31 | 0.66 | 0.59 | 0.41 | 0.52 |

**Table S2 Small simulation time results showing counterintuitive GWR efficiency over M-SVC.**

| Method | Time (sec) | Standard Deviation (SD) |
|---|---|---|
| **GWR** | 4.12 | 0.64 |
| **M-SVC** | 34.5 | 10.8 |
